# Automated artifact injection into sensing-capable brain modulation devices for neural-behavioral synchronization and the influence of device state

**DOI:** 10.1101/2023.07.31.23293393

**Authors:** Michaela E Alarie, Nicole R Provenza, Jeffrey A Herron, Wael F Asaad

## Abstract

**Background:** Sensing-enabled deep brain stimulation (DBS) devices enable opportunities to investigate correlations between neural activity and behavior. Unfortunately, these devices do not allow straightforward synchronization of neural data with external events.

**Objective:** To implement and assess an automated neural-behavioral synchronization system for a fully implanted DBS system.

**Methods and Results:** We describe a synchronization strategy that relies on computer-driven artifact injection via event-triggered transcutaneous stimulation (TS). We validated the temporal accuracy of the approach in two patients receiving DBS for treatment of Parkinson’s disease, observing consistently low jitter between task events and subsequent TS artifacts during DBS OFF (± 22.9ms) and ON (± 9.08ms) conditions. Notably, we observed that event-triggered TS was modulated by device state, where active circuitry during specific streaming modes influenced artifact injection in the data.

**Conclusion:** We describe a rigorous approach for neural-behavioral alignment using fully implanted DBS systems and demonstrate how accuracy of alignment depends on device state.

## Introduction

Deep brain stimulation (DBS) has increasingly been used for the treatment of various neurological and psychiatric disorders [1-2]. Sensing-enabled DBS devices allow chronic recording of ethologically relevant intracranial neural activity during concurrent stimulation [3-4]. A challenge that the field currently faces is how to leverage these neural recordings to identify biomarkers indicative of symptom states [2, 5]. Namely, there is still a need to define best practices with respect to synchronization with external data streams [6-7]. Unlike percutaneous research, where neural data synchronization with external events has been solved many times over, there are limited neural-behavioral synchronization methods for neural data recorded onboard fully implanted systems. Current strategies rely upon manually produced cross-channel artifacts to provide a means of time-locking data streams [8]. A rigorous approach requires an automated, precisely timed signal for reliable data alignment.

Here, we demonstrate the feasibility of event-triggered transcutaneous stimulation (TS) as a tool for injecting event-markers directly into recordings onboard sensing-enabled DBS platforms. First, we conducted benchtop testing to ensure consistently low latencies between task event markers and TS. We next validated our synchronization approach in humans, reporting on data collected onboard the commercially available Percept PC™ (Medtronic, Minneapolis, MN) in two patients with DBS of the subthalamic nucleus (STN) and globus pallidus internus (GPi) to treat Parkinson’s Disease (PD). We observed alignment performance is directly modulated by active device circuitry, namely streaming modes. Overall, this work improves current synchronization strategies onboard fully implanted DBS devices, permitting reliable observation of brain-behavior relationships.

## Materials and Methods

### Study Participants

Two patients implanted with Medtronic’s Percept PC™ to treat PD provided informed consent before participating in this study (Lifespan #1160521). DBS leads were implanted bilaterally in either the GPi (P1, Medtronic Sensight 33015) or STN (P2, Medtronic SenSight 33005), connected to one implantable pulse generator (IPG) located in the right chest.

### Experimental Design

Previous studies have aligned behavioral events with local field potentials (LFPs) via artifacts manually injected across recording streams from fully implanted and external systems [8]. We built on this approach, developing a computer-driven tool for injecting task events directly into device LFPs. Specifically, we configured a NeuroOmega recording system (Alpha Omega, Nazareth, Israel) as a triggerable TS platform (Fig. 1A). We programmed trial start markers within MonkeyLogic (https://github.com/michaela-alarie/Automated-neural-behavioral-alignment), a MATLAB (MathWorks, Natick, MA) application for creating and executing psychophysical experiments [9]. Trial start markers were sent from the task computer to the NeuroOmega system via a data acquisition interface (NIDAQ; BNC 2090A). We triggered TS pulses at the start of every trial using NeuroOmega functions within MonkeyLogic task code, sent via ethernet connection from the task computer to the NeuroOmega system. Both trial start event and stimulation times were logged by the NeuroOmega system.

**Figure 1:**
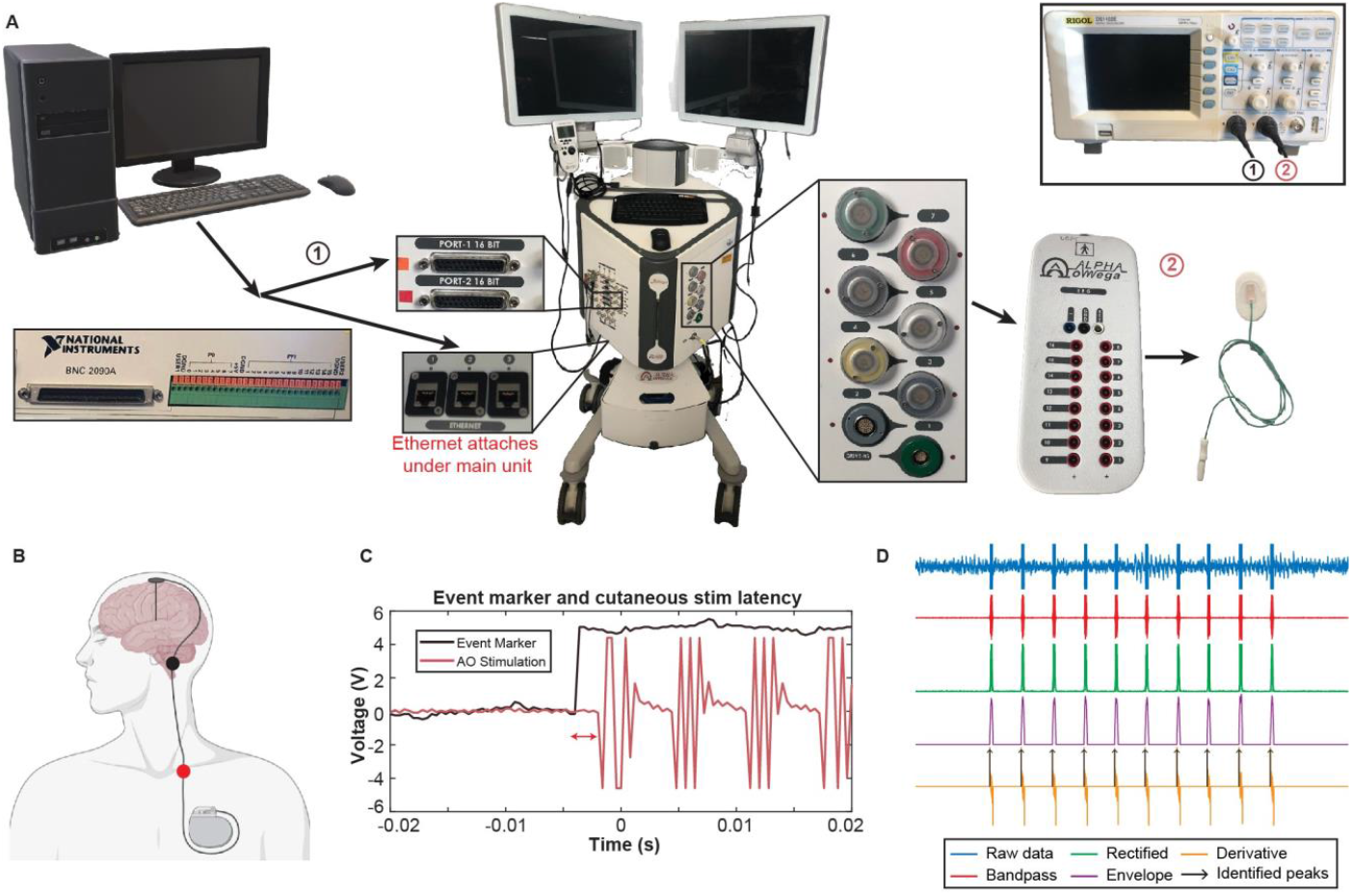
Experimental System Design. **A**: Experimental setup for delivering triggerable transcutaneous stimulation (TS). (1) and (2) are where oscilloscope probes measured latencies between event markers and stimulation. **B**: Location of surface contact placement (created with BioRender.com). The stimulation contact (red) was placed on the IPG-side clavicle and the reference contact (black) was placed on the IPG-side mastoid. **C**: Example visualization of latency testing with the oscilloscope (Pink: NeuroOmega system stimulation; Brown: event marker sent via DAQ). **D)** Neural data analysis steps for extracting artifact times including raw data (blue), bandpass filtered data (red), rectification (green), moving average and applied threshold (purple), derivative (yellow), and identified peaks (black arrows).

We delivered TS through two surface electrodes (Fig 1B; Neuroline 715, Ambu, Ballerup) placed on the mastoid (reference) and clavicle (stimulation) ipsilateral to the IPG [8]. Stimulation was transmitted through the EEG/EMG Headbox of the NeuroOmega system. Each TS burst had a duration of 0.5s, frequency of 80Hz, and pulse width of 0.5ms. Before initiating the task, we manually delivered TS pulses during *BrainSense*^*TM*^ *Streaming* at incremental amplitudes to assess the lowest TS amplitude for consistent artifact injection into device LFPs while maintaining patient comfort.

### Performance Verification

We conducted benchtop testing to measure latency within our computer-driven alignment system. We used an oscilloscope to measure the times of the event markers (N=50 trials; sent via NIDAQ analog output) and the subsequent TS pulses (Fig. 1A). Data was saved to a CSV file, where we computed latency as the time between task event markers and corresponding stimulation pulses (red arrows, Fig. 1C).

### Clinical Validation

We leveraged a sensing-enabled DBS platform to validate task-triggered TS alignment in two patients. In P1 we utilized Percept’s *BrainSense*^*TM*^ *Streaming* followed by *Indefinite Streaming* modes to compare alignment accuracies during DBS ON and OFF conditions, respectively. Because we did not observe TS artifacts during DBS OFF in P1, in P2 we evaluated task-triggered TS alignment during *BrainSense*^*TM*^ *Streaming* while setting DBS amplitude to 0mA. This condition, DBS Effectively OFF, allowed us to compare differences between active device circuitry across streaming modes. During *BrainSense*^*TM*^ *Streaming*, we recorded from contacts flanking the monopolar stimulation contact, producing 1 recording channel per hemisphere. During *Indefinite Streaming* we recorded from 3 channels per hemisphere.

### Data Analyses

We performed neural data analyses offline in MATLAB to identify times that TS artifacts appeared in the neural data (Fig. 1D). A 1^st^ order bandpass Butterworth filter (DBS ON: 70-90Hz; DBS OFF: 79.5-80.5Hz) was first applied to the data. We rectified the data by de-meaning and computing the absolute value. Next, we applied a moving average with a 50-sample sliding window. We then removed datapoints below 2x the mean to ensure only TS peaks were preserved. Next, we computed the second derivative to locate the initial times of each TS burst. We measured neural-behavioral alignment error as the time between NeuroOmega-logged TS transmission time and TS artifact times identified onboard device LFPs. Finally, identified TS artifacts were time shifted to ensure the mean event artifact alignment error was zero.

## Results

We measured the latency between task event markers and TS pulses using an oscilloscope as 2.57ms (mean) ± 2.88ms (standard deviation) (Fig. 2A). TS amplitude testing during *BrainSense*^*TM*^ *Streaming* ensured patient comfort and consistent artifact injection into the data with no discomfort reported. Final TS amplitudes were 1.4mA and 1.25mA for P1 and P2, respectively.

**Figure 2:**
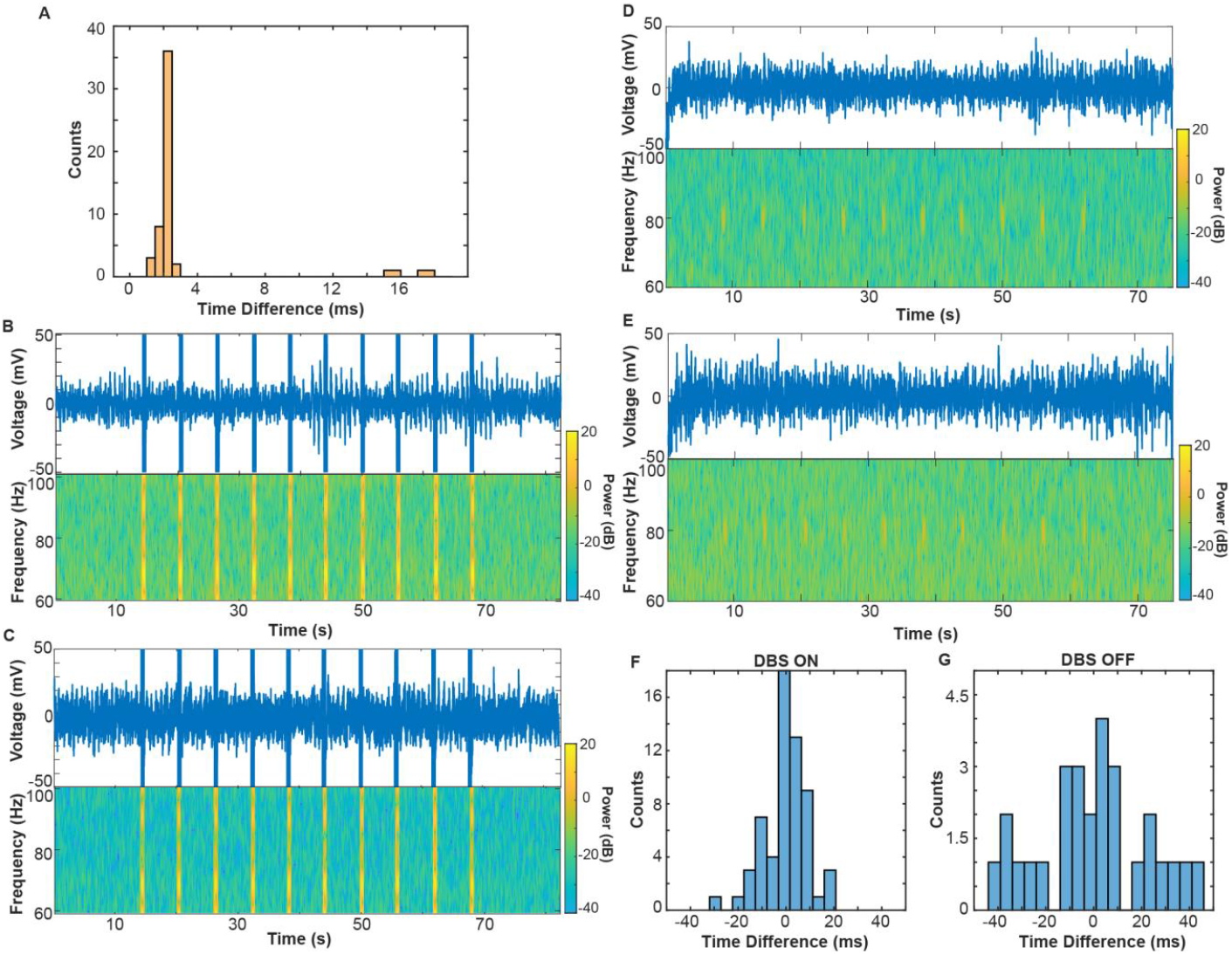
Benchtop and Clinical Validation of Event-Triggered Alignment. **A)** Time differences between task event marker and TS pulse sent from the NeuroOmega system. **B-C)** Example time domain (top) and spectrograms (bottom) for DBS ON condition on the left (B) and right (C) hemispheres from P2. **D-E)** Example time domain (top) and spectrograms (bottom) for DBS OFF condition on the left (D) and right (E) hemispheres from P2. **F-G)** Time differences between NeuroOmega-logged TS pulses and identified device LFP artifact times during DBS ON (F) and OFF (G) conditions across P1 and P2.

In both patients, the raw time domain data demonstrated distinct voltage increases during task-triggered TS during DBS ON (Fig. 2B-C), equal to the number of trials (top panels). Similarly, the time-frequency spectrograms (bottom panels) show full bandwidth power increases that align with time domain peaks.

In the DBS OFF condition (Fig. 2D-E), we observed no artifacts in the time domain data of either hemisphere (top panels). The time-frequency spectrograms revealed localized power increases at 80Hz (bottom panels) in P2, but we did not observe DBS OFF TS artifacts for P1. In DBS Effectively OFF, where the same device configuration was used for recordings as in the ON condition, we observed the same full bandwidth power increases as in the DBS ON condition.

Because there is no “ground truth” to assess the delay between the triggering of a TS pulse and the time at which it is recorded by the DBS pulse generator, accuracy of temporal alignment was measured as the relative jitter in TS pulse artifacts onboard the IPG compared to the jitter in the TS triggers in the NeuroOmega system. In the DBS ON (Fig. 2F; P1 and P2), OFF (Fig. 2G; P2), and Effectively OFF (P2) conditions, jitter was measured as ± 9.08ms, ± 22.9ms, and ± 12.5ms, respectively.

## Discussion

Synchronizing neural and task event markers is critical for ensuring that behavioral measures are accurately tied to neural features. Our work builds upon previously developed synchronization techniques [2, 9], describing an automated neural-behavioral alignment method for recordings onboard fully implanted DBS platforms.

Benchtop testing ensured TS was transmitted shortly after task event markers, where we measured consistently low latencies. We next validated our event-triggered TS alignment technique in vivo by calculating jitter between TS transmission logged by the NeuroOmega system and identified artifacts in the LFP data. Jitter during DBS ON and OFF conditions was measured under 50ms, which can be within the range required for analysis of many neural evoked response potentials [10]. Specifically, jitter appeared more localized in the DBS ON condition compared to DBS OFF condition (∼2x larger spread). This discrepancy is likely due to the increased prominence of the artifact in the neural data in the DBS ON condition. During DBS OFF, artifacts were localized to 80Hz, whereas full bandwidth artifacts were observed during DBS ON. We were able to identify these full bandwidth artifacts more accurately during post-processing, which enabled more accurate identification of TS onset times.

Inconsistent TS injection during the DBS OFF condition may be attributed to device circuitry changes across streaming modes. The case switches within the IPG are disconnected during *Indefinite Streaming*. This allows the IPG to be completely floating, permitting better rejection of external noise. Contrastingly, DBS Effectively OFF contains the same active circuitry as the DBS ON condition and therefore is more susceptible to noise, leading to similar jitter and artifact representations as in DBS ON. Our future work will explore TS configurations allowing more consistent artifact injection during *Indefinite Streaming*, including decreasing TS frequency, increasing TS amplitude, and shifting the reference contact from the mastoid to the top of the skull.

In sum, we implemented an event-triggered TS approach to synchronize external events with LFP recordings collected onboard fully implanted neuromodulation devices. Our approach eliminates the need for external recording streams to enable neural-behavioral alignment. Groups interested in implementing this technique might consider more economical and portable devices capable of event-triggered stimulation (i.e., the STG 4002 ®). This work is a step towards better synchronized neural recordings from sensing-enabled devices, leading to deeper insights into how neural features relate to symptom states.

## Data Availability

All data produced in the present study are available upon reasonable request to the authors

## CRediT authorship contribution statement

**Michaela E. Alarie:** Investigation, Software, Conceptualization, Writing – original draft. **Nicole R. Provenza:** Conceptualization, Writing – review & editing. **Jeffrey A. Herron:** Conceptualization, Writing – review & editing. **Wael F. Asaad:** Conceptualization, Writing – review & editing.

## Acknowledgements

We thank the participants for their involvement in the study as well as the Open Mind consortium for resources and guidance. Additionally, we thank Robert Raike, PhD, and the engineering team at Medtronic for guidance regarding TS sensing during Indefinite Streaming.

